# Cross-cohort reproducibility of AI-assisted nanopore methylation profiling establishes diagnostic equivalence to WHO-integrated diagnoses in adult CNS tumors

**DOI:** 10.64898/2026.11.26.26368828

**Authors:** Bernard S. Stikker, Susana Junqueira-Neto, Swip Draijer, Paul P. Eijk, Anabela Ferro, Jacqueline Egthuijsen, Mabel Pronk, Niels Verburg, Daoud Sie, Sybren L.N. Maas, Eelke Bos, Jorge Pinheiro, Rute Oliveira, Miguel Pinto, Jeroen de Ridder, Carlo Vermeulen, Pieter Wesseling, Jorge Lima, Bauke Ylstra, Evert-Jan Kooi

## Abstract

**Background:** Array-based methylation profiling with the Heidelberg classifier supports WHO-CNS5 classification of CNS tumors, but its substantial hands-on time, cost, centralized infrastructure and dependence on externally-hosted software limit broad implementation. Nanopore methylation sequencing combined with AI-based classification is a viable alternative, yet performance evidence required for implementation in adult CNS tumor diagnostics is limited. We test the equivalence of nanopore methylation profiling using the Sturgeon-v2 classifier against a WHO-CNS5-compliant integrated reference diagnosis incorporating array-based classification.

**Methods:** We compared Sturgeon-v2 classifications from nanopore sequencing of fresh-frozen tissue DNA with array-based reference diagnoses of matched FFPE tissue DNA, in two cohorts of 154 and 47 primary CNS tumors. Nanopore-derived copy-number alteration (CNA) profiles complemented Sturgeon-v2 predictions to resolve low-confidence calls. CNA-derived tumor cell fractions and histopathological evaluation determined tissue adequacy. Outcomes were categorized as concordant; discordant; additional analyses needed; or tissue inadequate for methylation profiling. Equivalence required the 95% upper confidence limit (95UCL) for discordance to be below 10%. Sturgeon-v2 classification and CNA analysis were implemented in the containerized pipeline NEURON.

**Results:** No discordant outcomes were observed (95UCL: 1.7%), with consistent performance across cohorts. Of 201 samples, 26 were inadequate due to low tumor purity or poor tissue quality. Of the remaining 175 samples, 167 were concordant with the reference diagnosis; eight required additional analyses.

**Conclusion:** Diagnostic equivalence and reproducibility across independent cohorts, together with the publicly available, locally-executable NEURON workflow, provide evidence and tools to support laboratory validation, accreditation and routine implementation of nanopore methylation profiling for adult CNS tumor classification.

**Key points:**

- Nanopore profiling showed diagnostic equivalence across two independent cohorts.
- NEURON automates local analysis supporting wider clinical implementation.
- Tissue quality and tumor cell fraction are essential for reliable classification

**Importance of the study:** DNA methylation profiling is valuable for CNS tumor diagnostics. However, the array-based technique relies on centralized capital- and labor-intensive infrastructure, limiting broader adoption. Validated alternative workflows are therefore needed to support clinical implementation across pathology laboratories, including resource-limited settings. Although nanopore methylation profiling has been shown to be feasible, performance and reproducibility evidence required for routine adult CNS tumor diagnostics has been limited. We demonstrate diagnostic equivalence of a nanopore-based workflow combining Sturgeon-v2 and CNA analysis against WHO-CNS5 integrated reference diagnoses across two independent cohorts, with no discordant outcomes. Our findings emphasize that tissue quality control, including assessment of tumor cell fraction, is essential for reliable classification. These results, together with the publicly available classifier running locally within our containerized pipeline NEURON, provide laboratories with evidence and tools for validation, accreditation and wider clinical implementation. Finally, our validation framework provides precedent for extending nanopore methylation profiling to other tumor entities.

## Introduction

Rapid advances in elucidating the (epi)genetic basis of primary central nervous system (CNS) tumors have yielded major biological and clinical insights[1–3]. Although histopathological evaluation has long formed the cornerstone of CNS tumor diagnosis, it does not have the capacity to adequately capture the extensive biological heterogeneity of these neoplasms while subject to substantial inter-observer variability. An early example was the incorporation of the 1p/19q status in IDH-mutant glioma to distinguish astrocytomas from oligodendrogliomas, thereby resolving the diagnostic ambiguity associated with the former oligoastrocytoma tumor type[4–6]. Molecular diagnostics have become central to CNS tumor classification, with new molecular insights continuously refining tumor taxonomy and clinical decision-making[1, 7]. This shift is reflected in the fifth edition of the World Health Organization (WHO) classification of CNS tumors (WHO CNS5), which mandates integration of histopathological and molecular features for diagnosis[8].

Next to molecular approaches such as next generation sequencing panels, genome-wide DNA methylation profiling has emerged as a particularly powerful diagnostic tool. WHO CNS5 strongly emphasizes its use as a desirable, and in some cases an essential criterion for accurate classification[9, 10]. Additionally, application of methylation profiling has enabled the identification of novel tumor entities, as reflected in the continuous expansion of the random forest–based Heidelberg CNS tumor classifier, which now distinguishes 184 methylation subclasses (v12.8)[1].

In routine adult neuro-oncology practice, array methylation profiling is frequently applied to adult-type diffuse gliomas, with data uploaded to the Heidelberg platform for classification[1]. This provides an objective classification while simultaneously yielding diagnostically relevant copy number aberrations (CNA) and O6-methylguanine-DNA methyltransferase (*MGMT*) promoter methylation status. Previous studies have shown that DNA methylation profiling confirms the original (immuno)histochemical diagnosis in most CNS tumors and provides diagnostically relevant refinement or reclassification of 10–25% of particularly of diagnostically challenging tumors[11–14]. However, array-based methylation profiling remains dependent on dedicated, capital-intensive microarray platforms, creating a need for diagnostically equivalent alternatives that require lower capital investment, offer greater workflow flexibility and allow local control over data analysis, thereby broadening access worldwide[15].

Recent advances in nanopore DNA sequencing offer a powerful alternative for molecular diagnostics. Low-coverage nanopore workflows coupled with machine-learning classifiers can rapidly achieve CNS tumor classification, including CNA profiles using sparse CpG methylation data[16–18]. Nanopore-based methylation and CNA profiling for CNS tumors have been validated in pediatric cohorts and used routinely in the intraoperative setting at several hospitals[19, 20]. In adults, feasibility has been shown[20], but the performance evidence needed for confident implementation and accreditation in routine diagnostics remains limited. Here, we formally test the diagnostic equivalence of a nanopore-based molecular diagnostic workflow on fresh frozen tissue (hereafter ‘nanopore workflow’), against strict WHO CNS5-compliant integrated reference diagnoses (hereafter ‘reference diagnosis’) that incorporated routine histomolecular diagnostics, including Heidelberg FFPE-based array classification, in two independent retrospective cohorts of predominantly adult-type diffuse gliomas. The nanopore workflow is implemented in NEURON[21], a locally executable, containerized pipeline that automates methylation-based tumor prediction with Sturgeon-v2 (Vermeulen et al. medRxiv, 2026), genome-wide CNA analysis and tumor purity quality control. The study is designed to reflect routine post-operative neuropathology practice and provides the performance evidence required for clinical implementation and accreditation.

## Methods

### Patient selection

We retrospectively assembled a Dutch cohort of 154 patients and Portuguese cohort of 47 patients, diagnosed with a primary CNS tumor, mostly comprising adult-type diffuse gliomas. In the Dutch cohort, eligible cases were selected based on prior DNA methylation profiling using the Illumina 850K EPIC or 900K EPICv2 array in combination with classification by the Heidelberg CNS tumor classifier (predominantly v11b4). To enable robust validation of the nanopore workflow against a reliable reference standard, the Dutch cohort only included cases with confident Heidelberg family scores (≥0.84 for v11b4 and/or ≥0.90 for v12.8) and fulfilling WHO CNS5 diagnostic criteria. Most of these cases (119/154; 77%) also achieved a confident Heidelberg class score. In addition, all reference diagnoses were independently confirmed using methylation data analysed with the NCI/Bethesda CNS tumor classifier v2.0[22]. In the Portuguese cohort, case selection was based on fulfilment of WHO CNS5 diagnostic criteria, rather than methylation classifier confidence alone, allowing assessment of the feasibility of nanopore sequencing in a diagnostic setting, without pre-selection of cases with high-confidence methylation-based classifiers outcomes. 26/47 samples had a confident Heidelberg v12.8 family score, of which 19 samples had a confident Heidelberg v12.8 class score as well.

Where available, additional molecular information, including immunohistochemistry and next-generation sequencing (NGS) gene panel data, was considered during case selection to support a confident reference diagnosis.

Oxford Nanopore Technology (ONT) sequencing currently requires fresh- or fresh frozen tissue-derived DNA to achieve optimal throughput and data quality, in contrast to methylation array workflows for CNS tumor diagnostics which are generally performed on FFPE-derived DNA. Consequently, only patients for whom matched frozen tumor tissue was available were included. This design resulted in a patient-matched rather than DNA-matched comparison between workflows, precluding a direct one-to-one DNA comparison. For all patients included, comprehensive histomolecular data were available (**Supplementary Table 1**).

### Tissue sectioning, DNA extraction and nanopore-sequencing

Samples for nanopore sequencing were processed as received from the neurosurgical procedure to reflect routine diagnostic practice, without preselection of tissue material. Representativity of frozen tumor material was assessed post nanopore sequencing, using haematoxylin and eosin (H&E) staining performed on 4 µm sections, obtained immediately before and after sectioning of tissue obtained for genomic DNA isolation.

For each fresh frozen tissue sample, 5 unstained sections of 20 µm thickness were collected for DNA extraction. In samples where the maximum tissue diameter was <3 mm, 10 unstained sections of 20 µm were collected. Genomic DNA was isolated using the QIAamp DNA Micro Kit (Qiagen; cat nr: 56304) according to the manufacturer’s instructions and eluted in 30 µL of nuclease-free water.

DNA quantity was determined using a Qubit 3 fluorometer (Invitrogen, Q33216) using the Qubit dsDNA BR assay kit (Invitrogen, Q33230). For DNA fragmentation, a solution of 250 ng genomic DNA in 50 µL nuclease-free water was sheared at 6000 x g for 1 minute using a Covaris G-TUBE (SKU: 520079) to achieve a targeted fragment size of approximately 5 kb after DNA library preparation. After fragmentation, DNA was concentrated using a ThermoSavant SpeedVac (SPD111V-115) for ∼30 minutes and reconstituted in 10 µL nuclease-free water. A total of 200 ng genomic DNA (in 10 µL) was required as input for library preparation with the ONT’s Rapid Barcoding Kit 24 (RBK114.24), following the manufacturer’s protocol.

In the Dutch cohort, samples were multiplexed with n=20 per PromethION flow cells (R10.4.1) and performed on an Oxford Nanopore Technologies’ P2 solo device for 48 hours (MinKNOW 25.03.9). In the Portuguese cohort, samples were multiplexed with n=5 using minION (R10) flowcells on a gridION device.

### Data processing and analysis

POD5 files generated by the Oxford Nanopore Technologies sequencer were processed on an HPC cluster using an in-house pipeline (https://gitlab.com/hg-aumc/bioinf/pipelines/sushi), using a GPU for basecalling and CPU for subsequent analyses. In short, basecalling was performed using Dorado (v0.7.2) with the high-accuracy model dna_r10.4.1_e8.2_400bps_hac@v4.2.0, enabling modified base detection for 5mC and 5hmC. Demultiplexing was performed using Dorado demux. Basecalled reads were aligned to the BSgenome.Hsapiens.NCBI.T2T.CHM13v2.0 reference genome using minimap2 (v2.26). Read-level quality control was conducted using NanoPlot and NanoLyse (v1.41.0), and summary metrics were compiled using MultiQC (v1.14).

Aligned bam files were sorted and indexed using Samtools (v1.18). Aligned bam files were analyzed using the NEURON pipeline[21], that performed Sturgeon prediction, CNA analysis, tumor purity estimation and reporting. In short, sturgeon is a CNS neural network classifier, originally based on the reference dataset published by Capper et al., 2018[11, 16]. Sturgeon-v2 has been trained on an extended dataset of nearly 10,000 methylation profiles and includes a redesigned classifier architecture (Vermeulen et al. medRxiv, 2026). CpG methylation calls were extracted using Modkit (v0.5.0, ONT). The resulting data were converted into the required input format for Sturgeon-v2 tumor prediction. Sturgeon-v2 predictions with a confidence score of ≥0.8 were considered confident. For all IDH-wildtype glioblastoma outcomes, ‘family’ scores are reported, which were obtained by summing the Sturgeon-v2 class scores, as it is currently only diagnostically relevant to diagnose a glioblastoma, rather than its molecular (sub)class (e.g. RTK1 or RTK2).

CNA analysis was performed using the QDNAseq package (v1.42.0), with a setting for sparse whole genome sequencing data by employing 500 kb bin sizes[23]. Read coverage was normalized against a 35x deep nanopore-sequenced Genome in a Bottle reference dataset[24] to correct for nanopore-specific mapping biases against the BSgenome.Hsapiens.NCBI.T2T.CHM13v2.0 genome. CGHcall (v2.68.0) was used to call CNAs[23, 25]. Bin annotations were generated using the BSgenome.Hsapiens.NCBI.T2T.CHM13v2.0 genome and were filtered to exclude low mappability regions, ENCODE blacklisted intervals, and bins with low complexity or insufficient base coverage[26]. Normalized read counts were segmented using the DNAcopy package (v1.80.0) to identify regions of genomic gain or loss[27]

Tumor purity and ploidy were inferred from the CNA data using ACE (v1.24.0)[28], applying the squaremodel function for model fitting. This approach infers absolute copy number states and estimates the tumor cell percentage based on best-fit integer copy number solutions across segmented regions. When the model estimates 100% tumor fraction, a visual inspection of fits and CNA profile were performed. If the profile was entirely flat, the estimate was adjusted to ‘below detection limit’. If not, the next best fit was used to estimate the tumor fraction. When ACE estimated the tumor purity below 20%, we deemed the CNA profile indicative of low tumor purity.

### Statistical analyses

For each case, Sturgeon-v2 prediction was followed by nanopore-derived CNA profiling to assign a final nanopore workflow outcome. Outcomes were categorized as concordant or discordant compared to the reference diagnosis; or additional analysis required or tissue inadequate for methylation profiling.

Samples were considered concordant only when the nanopore workflow yielded a WHO CNS5-compliant diagnosis, incorporating required defining molecular features such as *IDH* mutational status for astrocytoma and oligodendroglioma (the latter with 1p/19q codeletion) or class characteristic CNAs (gain of chromosome 7 combined with loss of chromosome 10 (+7/-10) and/or *EGFR* amplification) for glioblastoma, IDH-wildtype. Diagnostic equivalence was assessed by estimating the proportion of discordant samples across the combined cohorts. A one-sided 95% upper confidence limit (L) for this proportion was calculated using an exact binomial approach (Pearson-Clopper). Equivalence was defined *a priori* as L < 0.10.

Samples lacking a tractable nanopore workflow outcome, including low-confidence calls without low tumor purity estimates, or absence of required defining molecular features, were assigned the outcome ‘additional analysis needed’. Samples failing quality control due to poor tissue quality or low tumor purity, as determined by post-sequencing neuropathological review and tumor purity estimation inferred by ACE, were assigned the ‘tissue inadequate for methylation profiling’ outcome. ‘Additional analysis needed’ and ‘tissue inadequate for methylation profiling’ samples were not considered as true discordant in equivalence testing.

### Illumina EPICv2 arrays

To justify ‘tissue inadequate for methylation profiling’ and ‘additional analysis needed’ workflow outcomes, a subset of 23 DNAs of fresh frozen tissues, initially subjected to nanopore sequencing, underwent Illumina EPICv2 array profiling followed by Heidelberg v12.8 classification. As only one fresh frozen specimen was available per patient, this analysis provided information on the impact of intratumoral and sampling heterogeneity between the patient-matched samples and informed on whether Sturgeon-v2 low confident calls or control tissue predictions reflected insufficient tumor content of the fresh frozen specimens rather than true diagnostic disagreement. 200 ng DNA in 45 µL nuclease-free water was used as input material.

## Results

### General cohort characteristics

A total of 154 patients’ CNS tumors were included in the Dutch cohort with the following reference diagnoses: astrocytoma, IDH-mutant (n=38), oligodendroglioma, IDH-mutant, and 1p/19q-codeleted (n=35), and glioblastoma, IDH-wildtype (n=50). In addition, an ‘other CNS tumors’ category was included, consisting of meningioma (n=22), atypical teratoid rhabdoid tumor (n=1), ependymoma (n=1 spinal, n=1 myxopapillary and n=1 posterior fossa group B), high-grade astrocytoma with piloid features (n=1), a medulloblastoma, SHH-activated (n=1), pilocytic astrocytoma (n=2) and pleomorphic xanthoastrocytoma (n=1). The independent Portuguese cohort included glioblastoma, IDH-wildtype (n=43), astrocytoma, IDH-mutant (n=1), medulloblastoma, SHH-activated (n=1) and CNS tumor with BCOR internal tandem duplication (n=2). Detailed clinical, molecular, and radiological characteristics are provided in **Supplementary Table 2** for the Dutch cohort and **Supplementary Table 3** for the Portuguese cohort.

The results presented below follow the structure of nanopore workflow, starting with primary methylation-based tumor prediction by Sturgeon-v2, followed by assessment of CNA profiles, ultimately comparing the final workflow outcome to the reference diagnosis.

### Sturgeon-v2 performance

In the Dutch cohort, Sturgeon-v2 analysis yielded correct tumor predictions in 129/154 samples (83.8%) compared to the reference diagnosis (at the family level for glioblastoma, IDH-wildtype and class level for the rest of the entities). 124/154 samples (80.5%) exceeded the confidence threshold, while 5/154 samples (3.2%) were predicted correctly but did not reach the confidence threshold. 3/154 samples (1.9%) showed an incorrect prediction, one confident and two low-confident. In addition, 22/154 samples (14.3%) were predicted as control tissue, all with a confident prediction (**Figure 1**). Median confidence score was 0.99 (range: 0.47 – 1.0). Across the Portuguese cohort, Sturgeon-v2 yielded 40/47 (85.1%) confident correct predictions, 6/47 (12.8%) predictions of control tissue (four confident and two low-confident) and 1/47 (2.1%) low-confident incorrect prediction (**Figure 2**). Median confidence score was 0.99 (range: 0.48 – 1.0).

**Figure 1.**
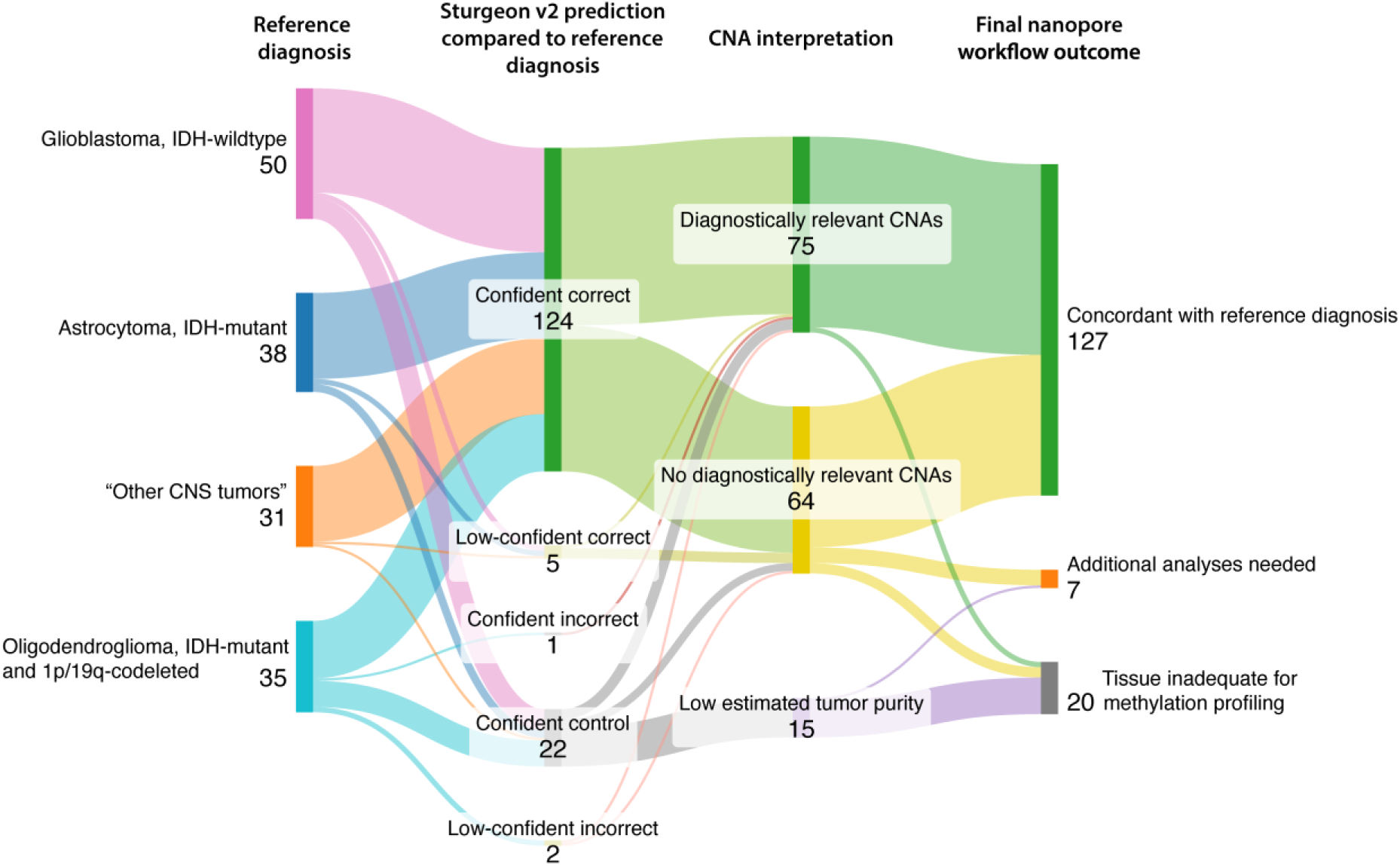
Sankey diagram showing the nanopore workflow for the Dutch cohort. Starting from the reference diagnosis, Sturgeon-v2 predictions are shown and subsequently, how nanopore-derived CNAs are interpreted towards the final nanopore workflow outcome. Sturgeon-v2 prediction and final nanopore workflow outcome are both compared to the reference diagnosis. CNA = Copy Number Aberration. Note: sample 108, which had a reference diagnosis of meningioma received a confident Sturgeon-v2 score of 1.00 for meningioma, but was retrospectively shown to consist of dura mater rather than tumor tissue, hence this sample flows from ‘confident correct’ to ‘tissue inadequate for methylation profiling’.

**Figure 2.**
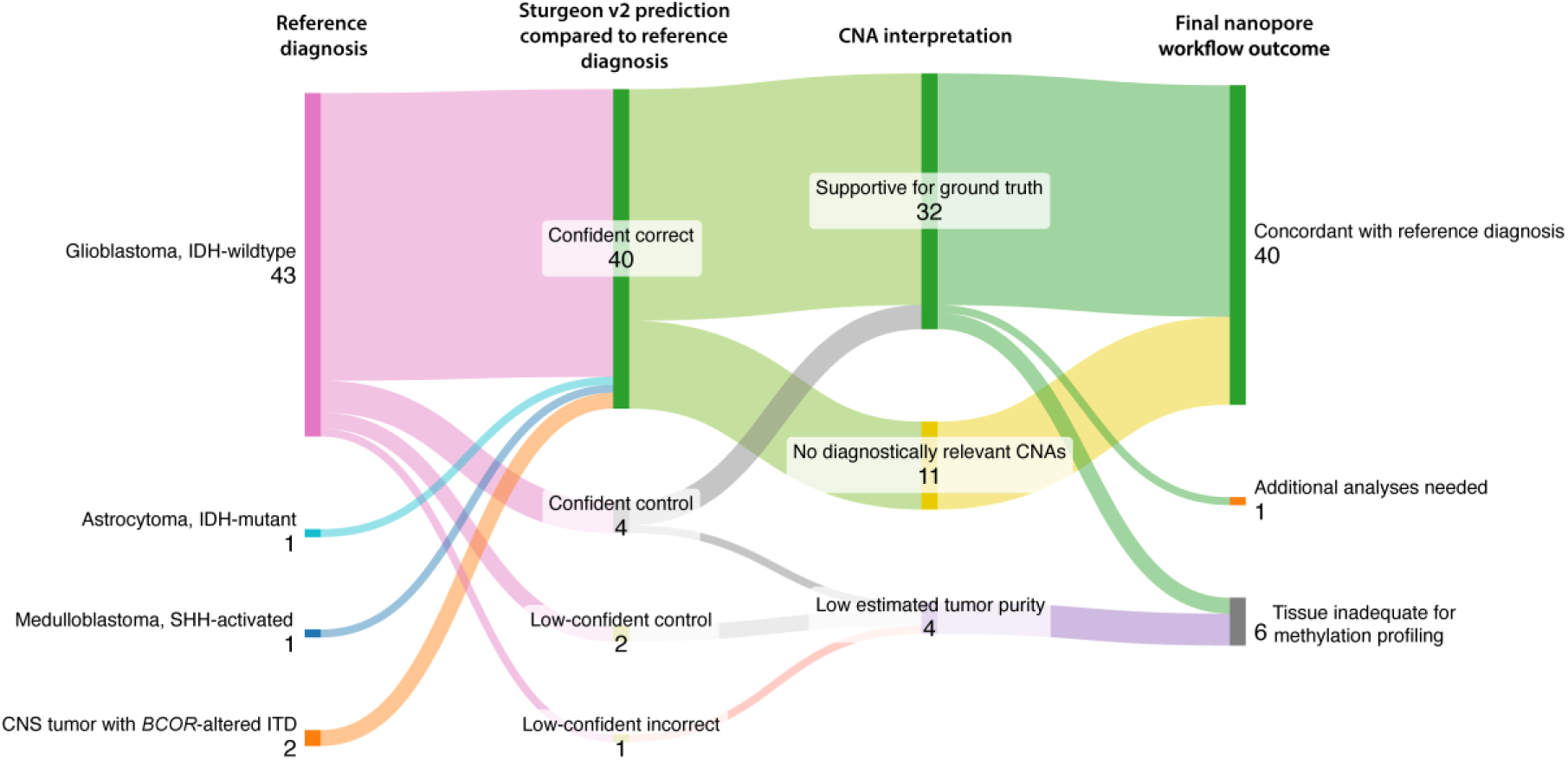
Sankey diagram showing the nanopore workflow in the Portuguese cohort. Starting from the reference diagnosis, Sturgeon-v2 predictions are shown and subsequently, how nanopore-derived CNAs are integrated towards the final nanopore workflow outcome. ‘Sturgeon-v2 prediction’ and ‘Final nanopore workflow outcome’ are both compared to the reference diagnosis. CNA = Copy Number Aberration.

The three incorrect predictions in the Dutch cohort were observed in the following samples; sample 109 was confidently predicted to be an astrocytoma, IDH-mutant (Sturgeon-v2 confidence score: 0.99), whereas the reference diagnosis was oligodendroglioma, IDH-mutant, and 1p/19q-codeleted (CNS WHO grade 3). Samples 103 and 122 were predicted with low-confidence to be a ganglioglioma (Sturgeon-v2 confidence score: 0.47) and an astrocytoma, IDH-mutant (Sturgeon-v2 confidence score: 0.56), respectively, whereas the reference diagnosis of both was oligodendroglioma, IDH-mutant, and 1p/19q-codeleted (CNS WHO grade 2). In the Portuguese cohort, sample IPATIMUP0067 was predicted subependymal giant cell astrocytoma (SEGA) with low confidence by Sturgeon-v2 (confidence score 0.77), while the reference diagnosis was an IDH-wildtype glioblastoma.

For these four samples, alternative CNS tumor classifiers (CrossNN and MethyLYZR) yielded both correct and incorrect predictions as summarized in patient-level heatmaps (**Supplementary Figure 1** and **Supplementary Figure 2**). All samples were further evaluated by integration of genome-wide CNA profiles as part of the nanopore workflow.

### Refinement of Sturgeon-v2 predictions following integration of CNAs

Nanopore-derived genome-wide CNA profiles were evaluated across all samples, focusing on predefined, entity-characteristic diagnostically relevant CNAs. Namely, +7/-10 and/or *EGFR* amplification in glioblastoma, IDH-wildtype, and complete 1p/19q codeletion in IDH-mutant oligodendroglioma. CNA profiles were not used to support diagnostic interpretation in astrocytoma, IDH-mutant, or in the ‘other CNS tumors’ category, as no defining CNAs are currently specified for these entities. In addition, tumor cell percentages were inferred from the CNA profiles using ACE as a quality control metric. Samples were categorized as having diagnostically relevant CNAs, no diagnostically relevant CNAs or low estimated tumor purity.

Across the Dutch cohort, 75/154 samples (48.7%) showed diagnostically relevant CNAs, 64/154 samples (41.6%) did not show diagnostically relevant CNAs, and 15/154 samples (9.7%) were suggestive of low tumor purity based on ACE (**Figure 1**). Between the Sturgeon-v2 confident correct predictions (124/154), CNAs were diagnostically supportive in 68/124 samples (54.8%) and did not show diagnostically relevant CNAs in 56/124 samples (45.2%). Between the low-confident correct calls (5/154), CNAs were diagnostically supportive in 1/5 samples (20.0%) and did not show diagnostically relevant CNAs in 4/5 samples (80%). Control tissue predictions (all confident; 22/154) were frequently associated with low estimated tumor purity. Specifically, 15/22 control tissue predictions (68.2%) showed low estimated tumor purity, while 4/22 (18.2%) showed diagnostically relevant CNAs and 3/22 (13.6%) did not.

Across the Portuguese cohort, 32/47 samples (68.1%) showed diagnostically relevant CNAs, while 11/47 (23.4%) did not show any diagnostically relevant CNAs. The remaining 4/47 samples (8.5%) had CNA profiles indicative of low tumor purity as estimated by ACE (**Figure 2**).

### Final nanopore workflow outcomes and diagnostic equivalence to the reference diagnosis

Across the Dutch cohort, 127/154 samples (82.5%) yielded a nanopore workflow outcome concordant with the reference diagnosis, 7/154 samples (4.5%) required additional analyses, and 20/154 samples (13%) were considered inadequate for methylation profiling by histopathological review and low-estimated tumor purity based on ACE. Excluding these QC failed samples, the workflow reached 94.8% concordance with the reference diagnosis (**Figure 1**). Finally, no discordant nanopore workflow outcomes were observed.

Across the Portuguese cohort, 40/47 samples (85.1%) yielded a nanopore workflow outcome concordant with the reference diagnosis, 1/47 samples (2.1%) needed additional analyses, and 6/47 samples (12.8%) were deemed inadequate for methylation profiling. Excluding these, the nanopore workflow reached 97.6% concordance with the reference diagnosis, while the remaining sample required additional analyses (**Figure 2**). Consistent with the Dutch cohort, no discordant nanopore workflow outcomes were observed in the validation cohort.

Across the combined cohorts, the 95% upper confidence limit (95UCL) for the true mismatch rate was 1.7%. To illustrate the diagnostic interpretation of the nanopore workflow, three cases are highlighted below.

### Exemplary samples illustrating the nanopore workflow

Sample 2 was selected as a clear example of a confident correct Sturgeon-v2 prediction (confidence score: 1.0). CNA analysis revealed diagnostically supportive *EGFR* amplification and +7/-10, with an inferred tumor purity estimate of 85%. Sturgeon-v2 based its prediction on 31,646 CpG methylation sites (**Figure 3A**). Of note, while part of the tissue was necrotic, Sturgeon-v2 was still able to correctly and confidently predict this sample, which emphasizes its sensitivity. The final workflow outcome was concordant with the reference diagnosis.

**Figure 3.**
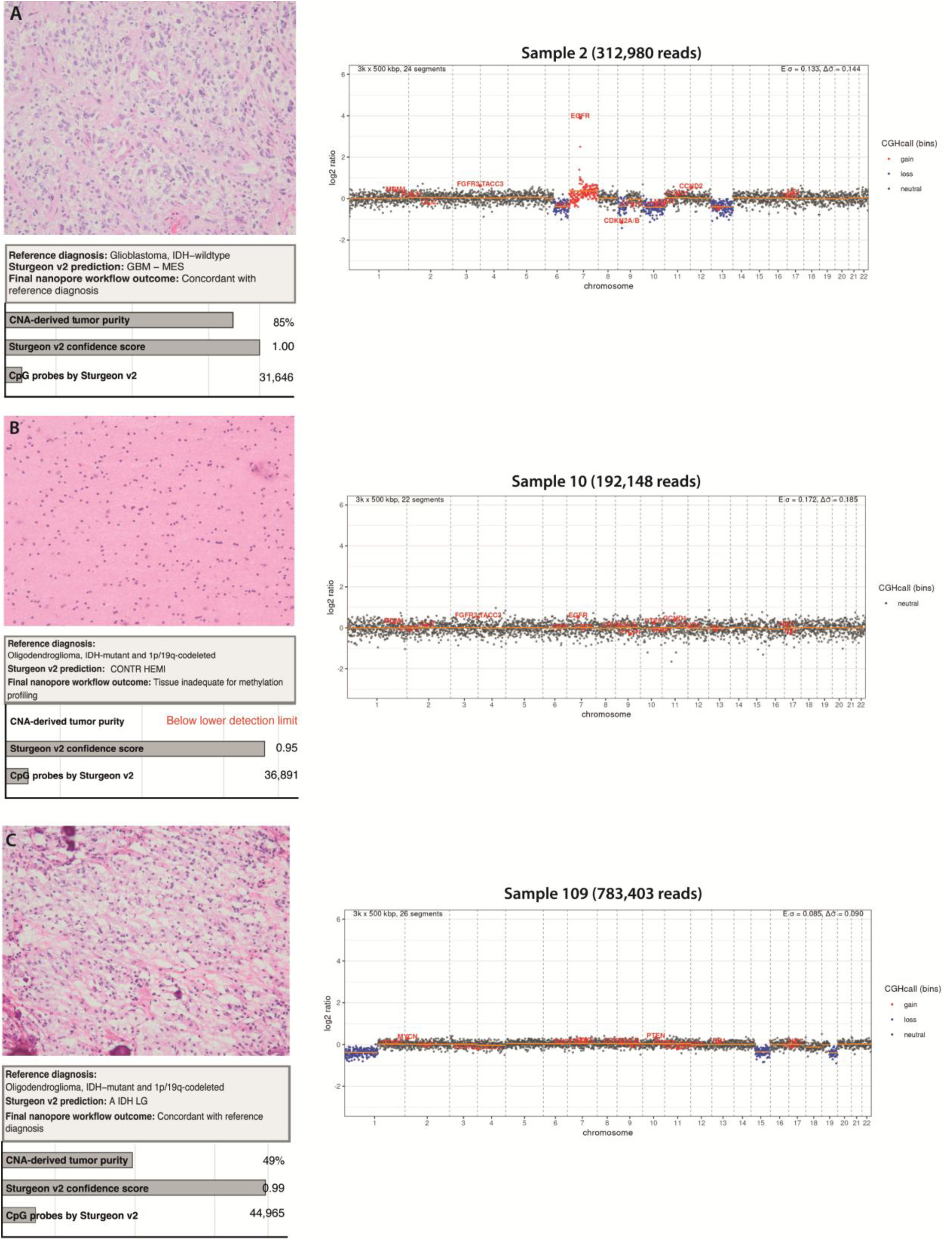
Representative samples are shown to illustrate the layers of the nanopore workflow at the individual-sample level. For each case, H&E–stained sections at 200x original magnification, nanopore-derived copy number profiles, and Sturgeon-v2 predictions are displayed. Sturgeon-v2 prediction and confidence score is shown, together with the number of CpG sites used for prediction and the ACE-based tumor purity estimate. CS = confidence score; H&E = haematoxylin and eosin.

Sample 10 was selected to illustrate a control tissue prediction with a high Sturgeon-v2 score (confidence score: 0.91) while the reference diagnosis was oligodendroglioma, IDH-mutant, and 1p/19q-codeleted (**Figure 3B**). ACE estimated a tumor purity below 20%, based on the CNA profile. Sturgeon-v2 used 36,891 CpG methylation sites for prediction. Reassuringly, rather than predicting a (low)-confident tumor, Sturgeon-v2 confidently predicted this sample as control tissue, emphasizing its specificity. DNA-matched analysis using Heidelberg v12.8 confirmed Sturgeon-v2 prediction (**Supplementary Figure 1**). Therefore, this frozen tissue sample was deemed inadequate for diagnostic methylation profiling.

Sample 109 was selected as it concerns the single confidently incorrect prediction by Sturgeon-v2, which predicted the sample to be an astrocytoma, IDH-mutant (confidence score: 0.99; **Figure 3C**). Sturgeon-v2 prediction used 44,965 CpG methylation sites, and the CNA-derived tumor purity estimation was 49%. The CNA profile showed a complete 1p/19q codeletion, which supports the oligodendroglioma reference diagnosis and resolves this sample towards a concordant final nanopore workflow outcome, as an astrocytoma, IDH-mutant cannot harbour a complete 1p/19q codeletion. Interestingly, Heidelberg v12.8 classification of this sample in a DNA-matched methylation array analysis showed subclassification of oligosarcoma (confidence score 0.90), despite a non-sarcomatous phenotype. In routine diagnostic practice, *bona fide* oligodendrogliomas, IDH-mutant and 1p/19q-codeleted may occasionally align with IDH-mutant astrocytoma on array-based methylation profiling, as previously described[29].

### DNA-matched array profiling for validation of selected Sturgeon-v2 outcomes

To validate control tissue and incorrect Sturgeon-v2 predictions (confident or low-confident), microarray profiling with Heidelberg v12.8 classification was performed on the same fresh-frozen DNA used for nanopore sequencing. This analysis was done to determine whether these predictions reflected insufficient tumor-derived methylation signal or were attributable to tissue heterogeneity (n=23; 17 confident control, 1 confident incorrect, 1 low-confidence incorrect, 2 confident correct, and 2 low-confidence correct Sturgeon-v2 predictions).

Overall, Sturgeon-v2 predictions and Heidelberg v12.8 classifications were concordant in 19/23 samples (82.6%). Among the 17 Sturgeon-v2 control tissue predictions, 15/17 (88.2%) were also independently classified as control tissue by Heidelberg v12.8. The remaining 2/17 samples (11.8%) were both confidently classified as glioblastoma, IDH-wildtype by the Heidelberg v12.8 classifier. 3/23 Sturgeon-v2 tumor predictions (13.0%) showed low confidence (two correct and one incorrect). Of these, Heidelberg v12.8 showed matching classifications in 2/3 samples (66.7%). The remaining low-confident incorrect sample was classified as low-confident correct by Heidelberg v12.8. The single confident incorrect Sturgeon-v2 prediction (sample 109) was correctly classified by Heidelberg v12.8 at the class level (confidence score 0.99), although incorrectly classified at the subclass level (oligosarcoma, IDH-mutant; confidence score 0.90) (**Supplementary Figure 3**).

### Heatmap highlighting patient-specific heterogeneity within tumor groups

To examine concordance and heterogeneity across classifiers and molecular features at the patient-level, an heatmap representation for the Dutch cohort is shown in **Figure 4**. Two versions of the Heidelberg classifier are shown, as a subset of samples was obtained after release of version v11b4 and could only be classified using version v12.8.

**Figure 4.**
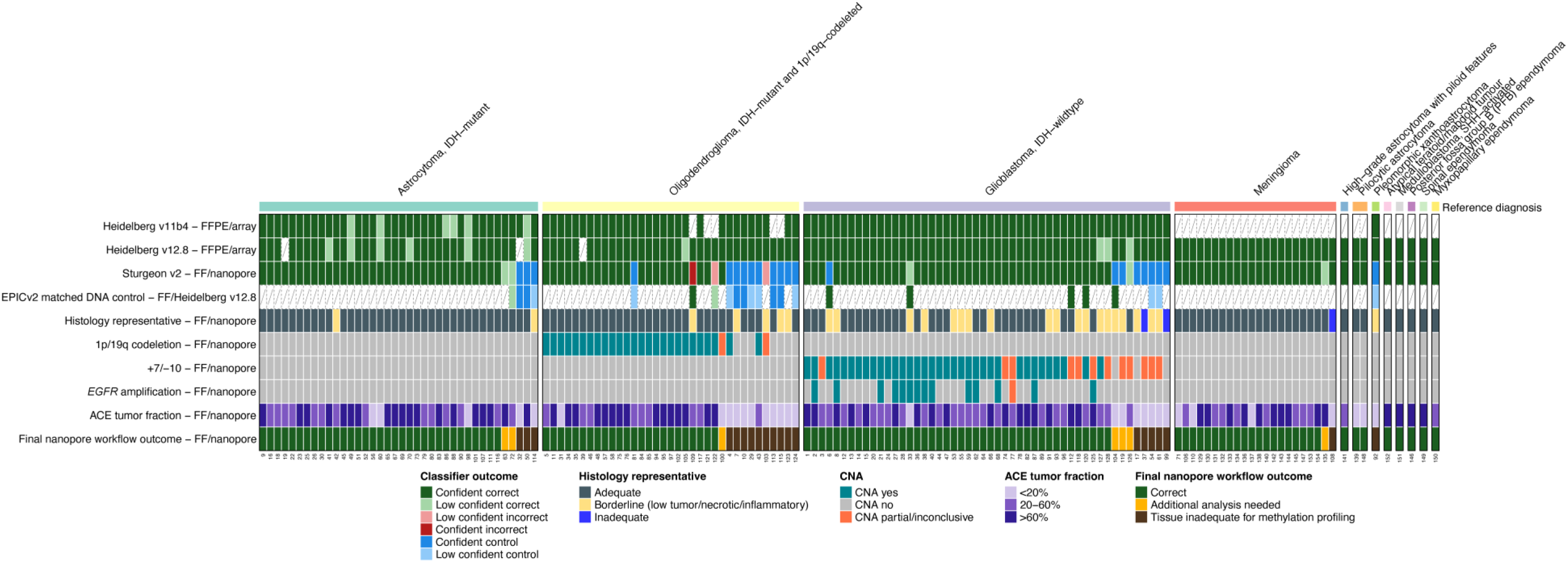
Patient-level heatmap showing classifier predictions, histological adequacy, copy number aberrations, tumor purity estimates, and final nanopore workflow outcomes for all samples in the Dutch cohort. Columns represent individual samples ordered by reference diagnosis; rows represent classifier outputs (Heidelberg v11b4, Heidelberg v12.8, Sturgeon-v2), histology, diagnostically relevant CNAs, ACE-derived tumor fraction, and final nanopore workflow outcome. FF = Fresh frozen; FFPE = formalin-fixed paraffin-embedded; CNA = Copy Number Aberration.

Across both cohorts, most confident correct Sturgeon-v2 predictions showed the expected patterns compared to the reference diagnosis, with CNA profiles, histology and tumor purity estimates aligning within individual samples. Additionally, low-confident tumor or control tissue predictions by Sturgeon-v2 were typically accompanied by low estimated tumor purity, and following histopathological review were subsequently assigned to ‘tissue inadequate for methylation profiling’. An extended heatmap including outcomes from CrossNN, methyLYZR and NCI/Bethesda classifiers, as well as additional CNAs is provided in **Supplementary Figure 1** for the Dutch cohort and **Supplementary Figure 2** for the Portuguese cohort.

### Locally executable workflow

The diagnostic equivalence and cross-cohort reproducibility presented here support implementation of the nanopore workflow in routine diagnostic practice. To facilitate local workflow implementation, we developed a containerized pipeline called NEURON that automates methylation extraction, Sturgeon classification, CNA analysis and tumor purity estimation from nanopore sequencing data and compiles this into one molecular report. To ensure reproducibility, both source code and container are publicly available through github and Zenodo (see *Data availability*).

## Discussion

DNA methylation profiling is nowadays a desirable and sometimes essential tool for the diagnosis of many CNS tumor entities[7, 9, 10] and incorporated into multiple diagnostic guidelines[30–32]. In this study, we demonstrated that a nanopore-based methylation sequencing workflow using Sturgeon-v2 and CNA analysis achieves diagnostic equivalence to a WHO CNS5-compliant integrated reference diagnosis for CNS tumors in adults. Together with the containerized pipeline NEURON and the publicly available Sturgeon-v2 classifier these findings support local implementation in routine post-operative neuropathology practice worldwide. No outcomes discordant with the reference diagnosis were observed in either the Dutch or the Portuguese cohort. After exclusion of samples inadequate for methylation profiling, concordance with the reference diagnosis reached 95.4% across the combined cohorts, with the remaining 4.6% requiring additional analyses. Compared with the workflow using the Sturgeon-v1 model[16], this performance reflects a shift toward increased specificity at the expense of modestly reduced sensitivity. Low-confidence predictions in otherwise adequate samples might relate to model recalibration following expansion of the training dataset[33]. In line with this, the number of confident incorrect predictions decreased from five with Sturgeon-v1 to one with Sturgeon-v2 (**Supplementary Figure 4**). Control tissue predictions and low-confidence calls were strongly associated with insufficient tumor signal, typically due to low tumor cell content or extensive necrosis. This was supported by complimentary microarray-based analysis using the same DNA as used for nanopore sequencing, where most Sturgeon-v2 control tissue predictions were validated by control tissue classifications using the Heidelberg v12.8 classifier (**Supplementary Figure 3**). These findings underscore that most discrepancies do not arise from incorrect prediction, but from biological and technical limitations such as low tumor purity or extensive tissue necrosis, observed in both the Dutch and Portuguese cohorts. Similarly, samples requiring additional analyses frequently showed only partial or absent glioblastoma-associated CNAs (+7/−10 and/or *EGFR* amplification), aberrations that are supportive yet do not independently establish a glioblastoma diagnosis[7, 34].

Applying an ACE-estimated tumor purity cut-off of 20% after nanopore sequencing excluded 18/22 (81.8%) control tissue predictions of Sturgeon-v2 in the retrospective setting (**Supplementary Figure 5**). The increased number of Sturgeon-v2 control tissue predictions than expected from the reference diagnoses likely reflects differences in selection of material from the patients following the procedures of formalin fixation versus fresh material. For routine methylation array workflows, pathologists typically select the most representative tumor tissue from multiple FFPE blocks. In some cases, a representative region within the selected tissue is then demarcated on an FFPE section prior to DNA isolation which ensures optimal tissue quality and high tumor cell percentage. In contrast, nanopore sequencing in this study was performed using a single fresh-frozen tissue sample without prior tumor region selection, making the workflow more susceptible to low tumor purity and tissue necrosis. For clinical implementation, we recommend to mitigate this limitation before nanopore sequencing through careful sampling of representative tumor regions and, where feasible, inclusion of multiple fresh tissue fragments to select the most representative tissue.

Importantly, there were still occurrences where Sturgeon-v2 prediction was either inconclusive or confidently wrong, even with sufficient tumor purity. Integrating CNA data proved crucial in resolving those cases, as exemplified by confident misclassification of Dutch sample 109 (**Figure 3C**). Thus, confident predictions should not be accepted uncritically, as rare but relevant misclassifications occur. Conversely, low-confidence results should not be dismissed when in line with other data, as high-confidence scores are achieved in only 50–70% of cases in real-world cohorts, indicating that low-confidence predictions remain common but informative[13, 14, 35, 36].

From a technical perspective, multiplexing up to 20 samples (dependent on the type of flow cell) and sequencing for 48 hours provided sufficient coverage for robust classification and CNA profiling in most cases. Increasing sequencing depth by reducing multiplexing and increasing sequencing time would be expected to further improve CNA resolution and can enable more reliable detection of additional genomic features, including somatic mutations, structural variants such as translocations and *MGMT* promoter methylation [20, 37, 38]. However, this must be balanced against cost-efficiency in routine practice.

Several limitations should be considered. While *MGMT* promoter methylation assessment is feasible[20, 37, 39], this remains dependent on sufficient CpG promoter coverage and was not addressed in this study. Nevertheless, NEURON is equipped to predict *MGMT* promoter methylation status by leveraging a previously published prediction model[40]. In addition, the requirement for fresh frozen tissue represents a logistical challenge, as FFPE material remains the standard in diagnostic pathology. On the other hand, nanopore sequencing measures methylation directly on native DNA, thereby avoiding bisulphite conversion artifacts[41]. Nevertheless, recent technological advances may enable high-quality, high-throughput analysis of FFPE-derived DNA, facilitating simultaneous genome-wide methylation profiling and whole-genome sequencing in routine pathology workflows (Kokoris et al. bioRxiv, 2026). Informative methylation profiles can also be generated from FFPE material using nanopore sequencing; however, fragmentation and chemical damage of FFPE-derived DNA can increase variability in sequencing output and may impact cost and time required for robust classification[42, 43]. In parallel, nanopore whole-genome sequencing and targeted enrichment approaches such as adaptive sampling may further expand the diagnostic capabilities of the proposed workflow by improving coverage of clinically relevant genomic regions[39]. Furthermore, although our cohort included several uncommon CNS tumor entities, it was primarily powered to evaluate the most prevalent CNS tumors in adults encountered in routine diagnostics. Consequently, performance estimates for rarer entities remain limited. However, as the Sturgeon-v2 model was trained on a broad reference dataset encompassing many rare tumor entities, this limitation relates to clinical validation rather than classifier development.

In conclusion, the nanopore workflow demonstrated diagnostic equivalence to the WHO CNS5-compliant integrated reference diagnosis for CNS tumors in adults, which incorporated conventional array-based classification. Integration of genome-wide CNA profiling and tumor purity assessment proved essential for robust diagnostic interpretation, particularly in low tumor purity samples. These findings provide the necessary performance evidence supporting implementation of nanopore methylation sequencing using Sturgeon-v2, as a scalable and potentially cost-effective alternative to array-based methylation profiling for routine CNS tumor diagnostics in adults. Our containerized pipeline NEURON facilitates a locally executable and reproducible workflow, enabling implementation across pathology laboratories worldwide. As methylation-based classifiers expand, this study may serve as precedent towards extending the approach to other tumor types, particularly for diagnostically challenging entities such as head and neck including salivary gland tumors, lymphomas, and sarcomas, where methylation profiling is promising[44–46].

## Required statements

### Ethics

The research was approved by the Medical Ethics committee of Amsterdam University Medical Center (METC 2024.0773). All included patients provided written informed consent for participation in the biobank.

### Authorship

B.S. designed and conducted Dutch cohort experiments, performed bioinformatic analyses on all data and wrote the manuscript. S.N., R.O. and A.F., designed and conducted Portuguese cohort experments. S.D. provided clinical implementation expertise and aided in study discussions. M.Pi. performed bioinformatics analyses of Portuguese cohort. J.L. aided in Portuguese cohort coordination, experimental design and data analysis. J.P. identification and pathological assessment of Portuguese cohort. J.E. and M.Pr. and aided in setting up future diagnostic runs, writing and validating run protocols. P.E. provided technical assistance in nanopore experiments. N.V. and E.M.B. collected study material. D.S. aided in study discussions and facilitated HPC use and P2solo runs. S.L.N.M. provided non-glioma material. C.V. aided in study conceptualizing, technical expertise and study discussions. J.d.R. aided in study conceptualizing. P.W. aided in study conceptualizing, study discussions and neuropathological expertise. B.Y. aided in study conceptualizing, study discussions, manuscript writing and technical expertise. E.J.K. aided in study conceptualizing, experiment design, study discussions, neuropathological expertise, pathological assessment and coordination of the Dutch cohort, and manuscript writing. All authors contributed to manuscript revisions.

## Data Availability

Microarray and nanopore data are available in EGA (EGAS00001008560). NEURON is available at <u>tgac-vumc/NEURON</u> and archived at Zenodo (https://doi.org/10.5281/zenodo.24335955). The Apptainer container is available at https://doi.org/10.5281/zenodo.24336315

## Acknowledgements

We would like to express our gratitude towards Santiago Bolaños Meneses for data management in this project. In addition, we’d like to extend this to the Helios HPC support team and the Core Facility Genomics from Amsterdam UMC for their continued support in facilitation of data generation, analysis, and software maintenance in particular Roxane Boyer, Martin Haagmans, Lars Nooi, Naomi van Es, Karima Hajo and Reinier Vleugels.

---

**Supplementary Figure 1.**
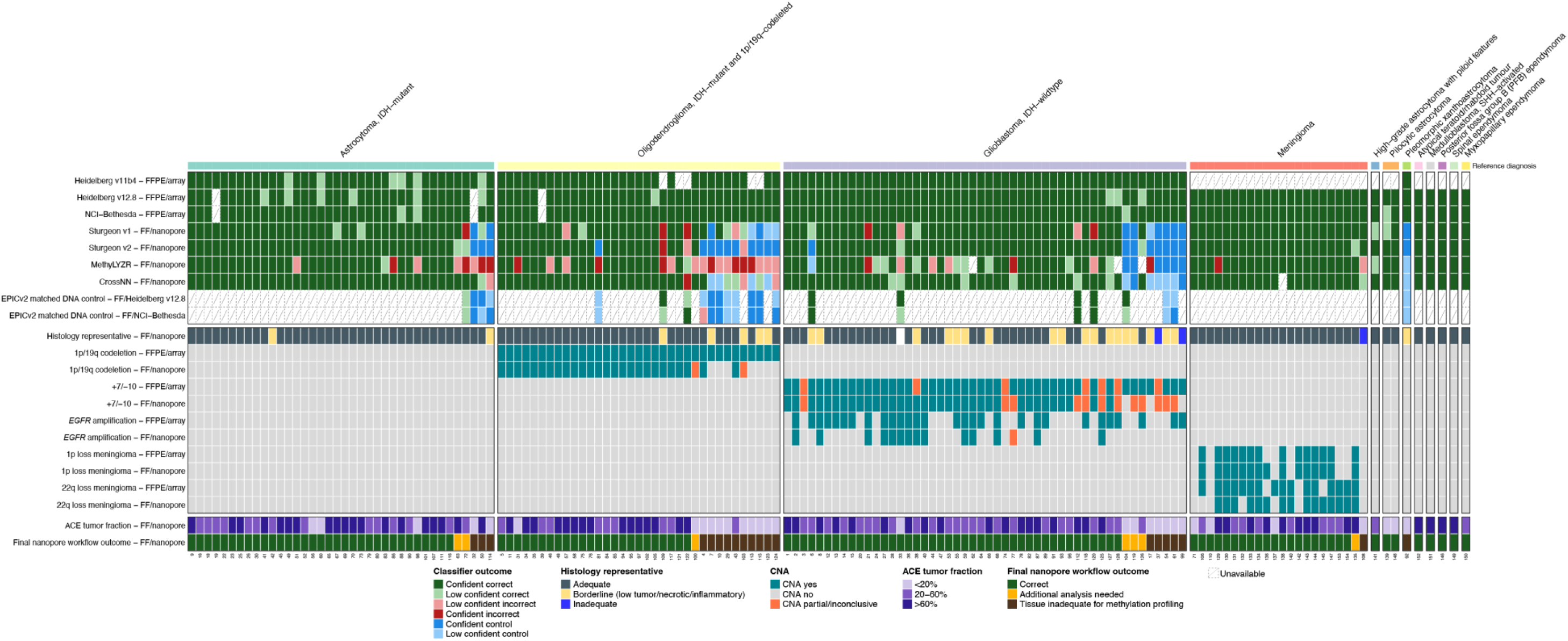
Extended heatmap overview of the Dutch cohort. Top rows include different CNS tumor classifiers ran on both array data (FFPE) and patient-matched nanopore data (FF). DNA matched EPICv2 controls are shown for both Heidelberg v12.8 and NCI/Bethesda classifiers. Tissue representativity is classified into adequate, borderline and inadequate based on histopathological assessment on H&E. Characteristic CNAs assessed are shown for both FFPE/array and FF/nanopore and include complete 1p/19q codeletion, +7-10, EGFR amplification, and 1p/22q loss for meningiomas. ACE-estimated tumor purity is shown for the FF/nanopore samples. Finally, the final nanopore workflow outcome is shown. Samples 62, 126 and 128 did not yield a technically sound outcome for MethyLYZR. Sample 138 did not yield a tractable outcome from CrossNN. FF = fresh frozen; FFPE = formalin-fixed paraffin-embedded; TCP = Tumor Cell Percentage; H&E = Haematoxylin & Eosin.

**Supplementary Figure 2.**
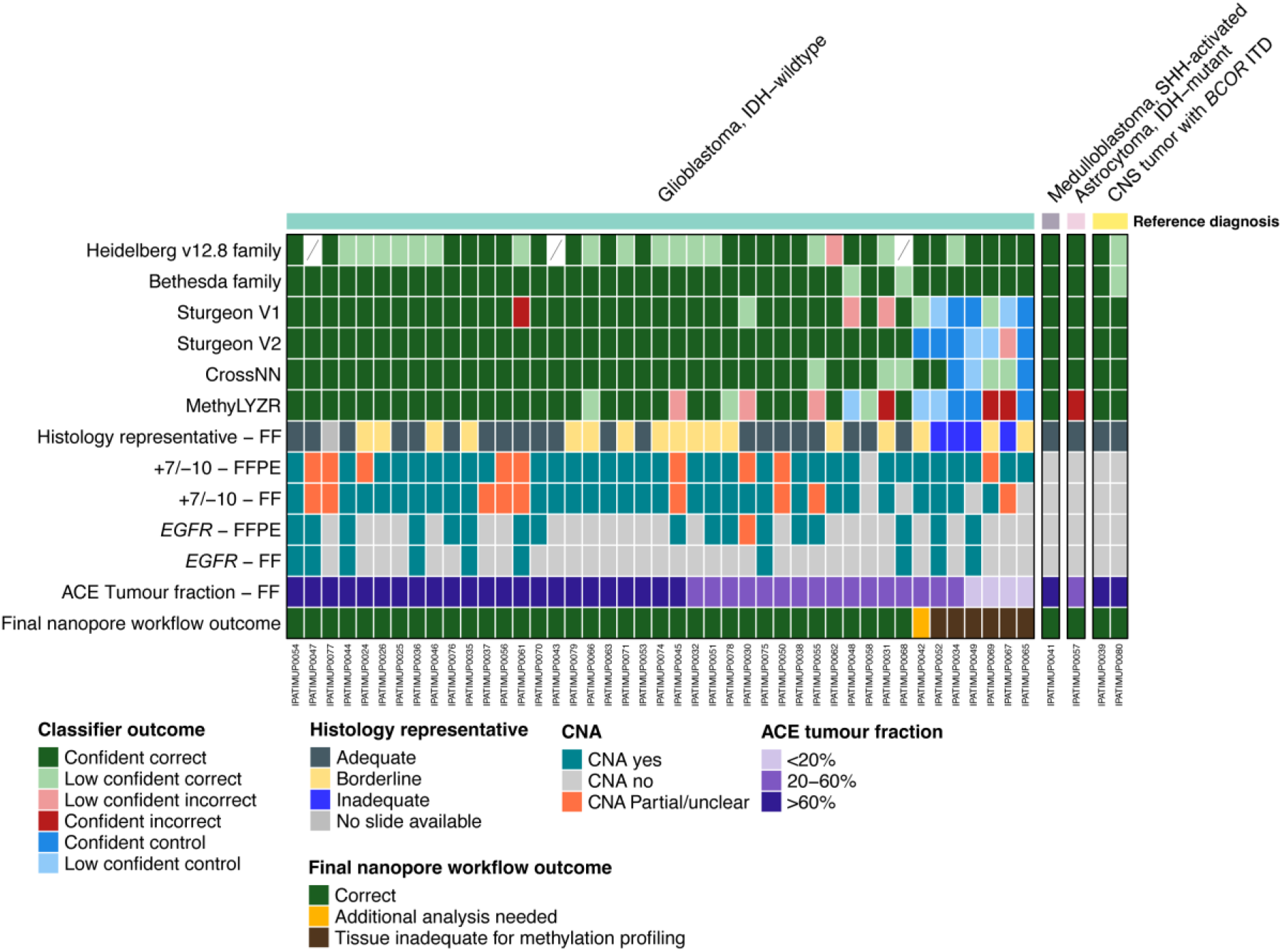
Heatmap overview of the Portuguese cohort. The top 6 rows show different CNS tumor classifier predictions, followed by H&E representativity assessment of the FF material. The next 4 rows show the presence of diagnostically relevant CNAs followed by CNA-inferred tumor cell percentage by ACE. The lower row shows the final nanopore workflow outcome. FF = fresh frozen; FFPE = formalin-fixed paraffin-embedded; TCP = Tumor Cell Percentage; H&E = Haematoxylin & Eosin; CAN = Copy Number Aberration.

**Supplementary Figure 3.**
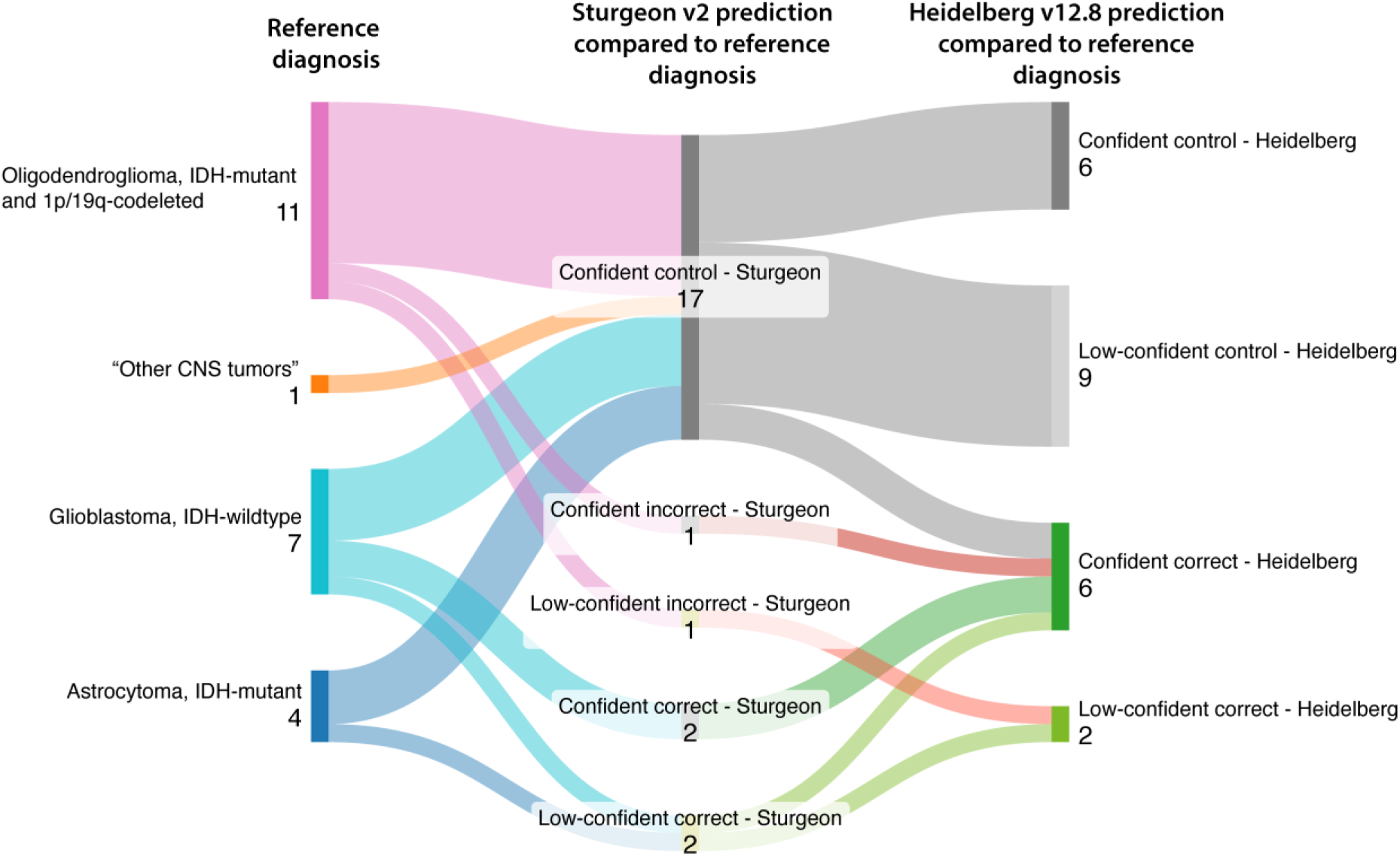
Sankey overview for the DNA-matched EPICv2-array analysis, showing the reference diagnosis, Sturgeon-v2 prediction and Heidelberg v12.8 classification. Note that the majority of Sturgeon-v2 control tissue predictions are validated by Heidelberg v12.8 outcomes.

**Supplementary Figure 4.**
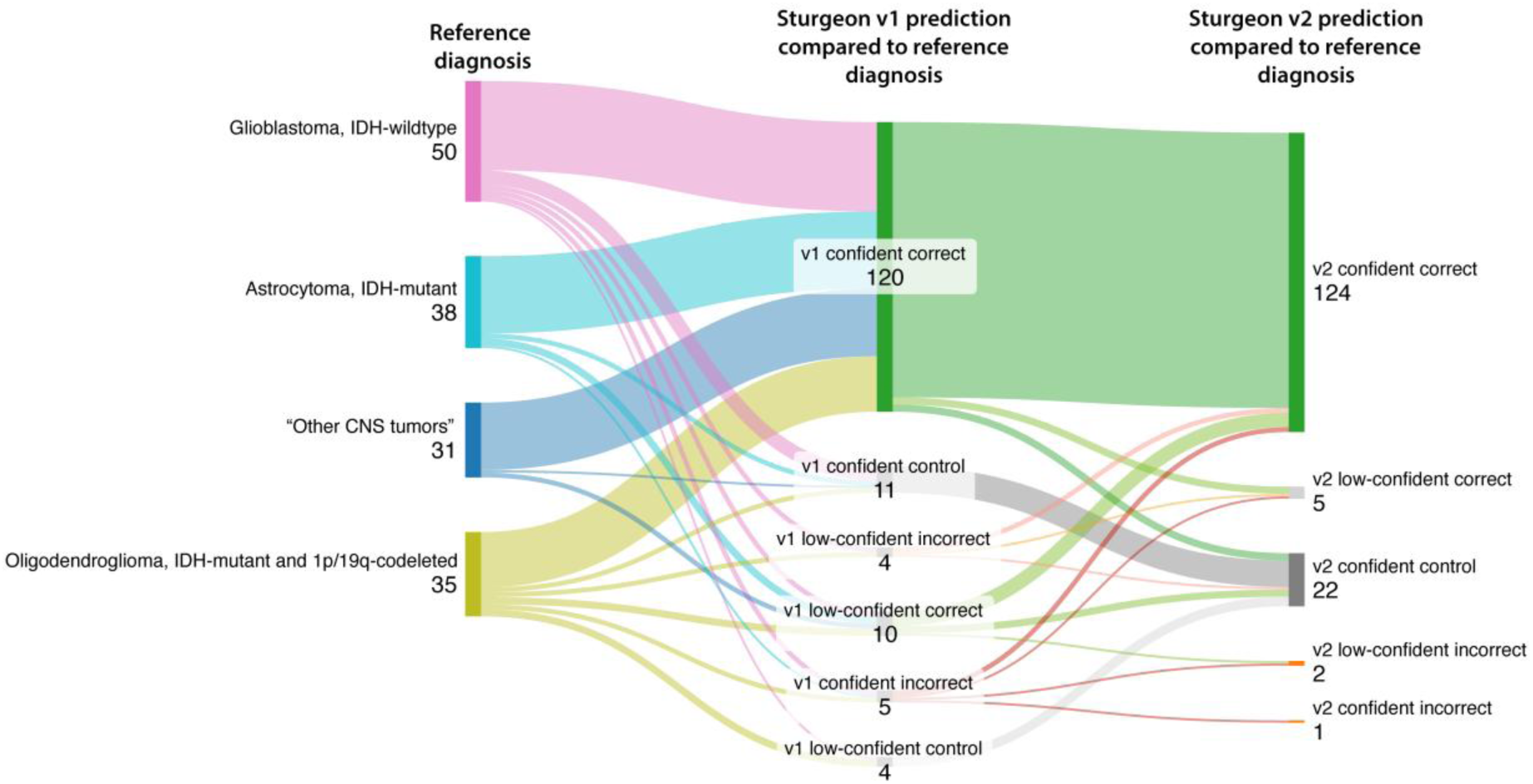
Sankey overview comparing Sturgeon-v1 and Sturgeon-v2 predictions for the Dutch cohort. CNA interpretation has not been taken into account. Sturgeon-v1 and v2 show comparable results. However, Sturgeon-v1 shows higher sensitivity (although with lower confidence scores) compared to Sturgeon-v2. In contrast, Sturgeon-v1 shows higher specificity with markedly less incorrect predictions (5 versus 1).

**Supplementary Figure 5.**
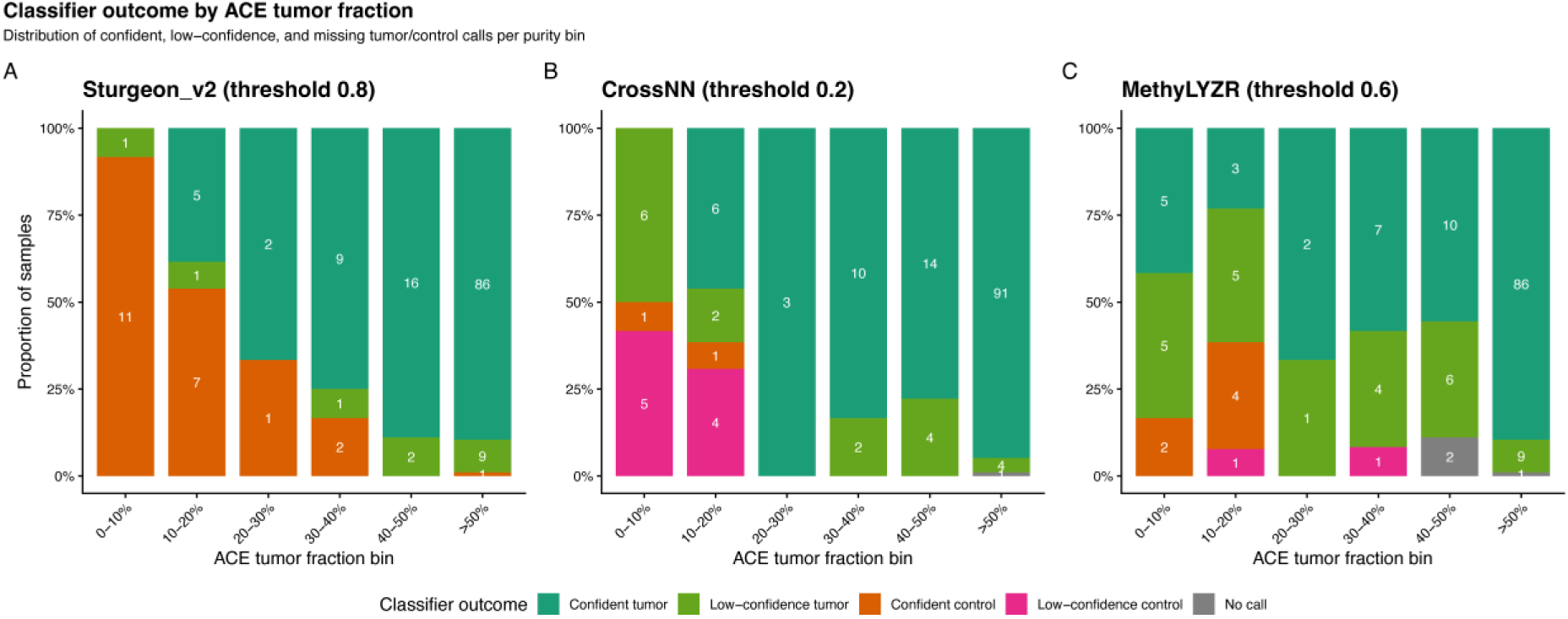
**A**) Stacked bargraph showing CNA-derived tumor purity by ACE, against Sturgeon-v2 predictions. **B**) Same as panel A but for MethyLYZR outcomes. **C**) Same as panel A but for CrossNN outcomes. NA = no classifier outcome available.

